# Calibrated Functional Data Decreases Clinical Uncertainty for *KCNH2*-related Long QT Syndrome

**DOI:** 10.1101/2025.02.05.25321617

**Authors:** Chai-Ann Ng, Matthew J. O’Neill, Samskruthi R. Padigepati, Yi-Lee Ting, Flavia M. Facio, Matteo Vatta, Sarah R. Poll, Jason Reuter, Jamie I. Vandenberg, Brett M. Kroncke

## Abstract

Rare missense variants are often classified as variants of uncertain significance (VUS) due to insufficient evidence for classification. These ambiguous findings create anxiety and frequently lead to inappropriate workup, colloquially referred to as the ‘diagnostic odyssey’. Well-validated high-throughput experimental data have the potential to significantly reduce the number of VUS identified by clinical genetic testing, though the extent of this reduction and the optimal strategies to achieve it remain unclear.^1^

---

Long QT Syndrome (LQTS) is a Tier 1 monogenic condition, with ~40% of cases arising from loss-of-function variants in *KCNH2*. Sentinel manifestations include sudden cardiac death. Pre-emptive genetic testing allows for proactive clinical management in people with identified pathogenic variants. To date, Invitae Corporation (now part of Labcorp) has reported over 1,900 unique variants in *KCNH2* in over 190,000 patients, which are submitted to ClinVar on a routine basis. Our group recently calibrated two functional assays to support variant classification and stratify risk: a Multiplexed Assay of Variant Effect (MAVE) (Figure [A]) and Automated Patch Clamp (APC) (Figure [B]) assay.^2^ In this study, we tested the value of functional data, when added to a validated point-based variant classification framework (Sherloc) (Figure [C])^3^, in reducing uncertainty in a real-world clinical genetics setting.

**Figure.**
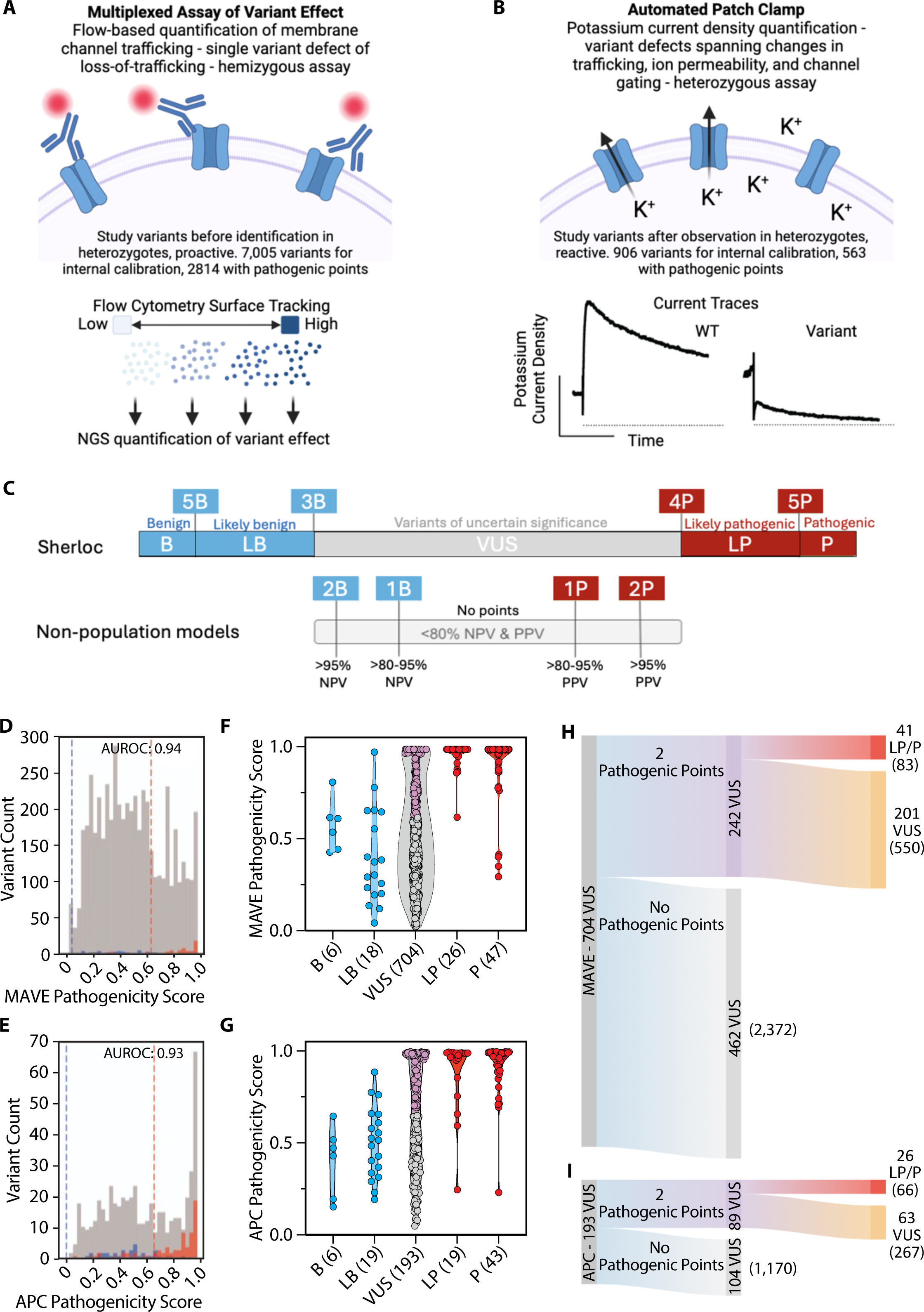
Effectiveness of Multiplexed Assay of Variant Effect (MAVE) and Automated Patch Clamp (APC) data for VUS reclassification in *KCNH2-*related LQTS. (**A**) The *KCNH2-*MAVE used fluorescence-activated cell sorting and Next Generation Sequencing (NGS) of HEK cells to quantify variant membrane abundance compared to that of WT. (**B**) APC quantified potassium current density, which captured variant effects on ion permeability and channel gating, in addition to trafficking deficiencies. Both assays studied stably transfected HEK293 cells. The published MAVE and APC datasets were used as input to model negative predictive value (NPV) and positive predictive value (PPV). (**C**) Benign (B) or Pathogenic (P) point thresholds for variant classification. Models based on functional evidence can contribute up to 2 benign or 2 pathogenic points depending on NPV and PPV. Establishment of PPV and NPV using benign and pathogenic *KCNH2* labels with (**D**) Histogram of pathogenicity scores for MAVE data and (**E**) APC data. Pathogenic and benign labels are coloured red and blue, respectively. VUS are coloured grey. Variants are classified in Sherloc based on the aggregate score of all pathogenic evidence or the aggregate score of all benign evidence, evaluated independently. Classification as of 6 May 2024 for variants with corresponding MAVE data (**F**) or APC data (**G**). Likely pathogenic (LP) or pathogenic (P) variants are red, likely benign (LB) or benign (B) variants are blue, and variant of uncertain significance (VUS) are grey. VUS with eligible pathogenic points are violet. The number of variants is indicated in brackets. Sankey plots show the reassessment of 704 or 193 VUS with pathogenic points from MAVE (**H**) or APC (**I**), respectively. The number of patients is indicated in brackets.

A total of 7,005 MAVE and 906 APC variants were calibrated against a set of internally curated 50 or 38 benign and 216 or 168 pathogenic variant controls for MAVE and APC (Figure [D-E]), respectively; the larger number of MAVE variants reflects its higher throughput and lower cost per data point compared to APC. The AUROC of both datasets was ≥0.8 (Figure [D-E]), passing internal required standards for performance, and allowing the data to be integrated into Sherloc based on negative predictive and positive predictive values (NPV, PPV) (Figure [C])^4^. Based on the PPV threshold that discriminates pathogenic variant controls, pathogenic points could be assigned to 2,814 (MAVE) and 563 (APC) variants respectively. No benign points could currently be assigned to either dataset due to an insufficient number of benign missense controls to establish a robust NPV.

We evaluated the effectiveness of incorporating this validated functional data in classifying missense VUS in the dataset up to May 2024. Data were deidentified and approved for use in this study by WCG Institutional Review Board (study number 1167406); these data are available from the corresponding author upon reasonable request. Figures [F-G] show the latest classification of variants with corresponding MAVE and APC data. Of the 704 missense VUS reported in 2,993 individuals with MAVE data, 242 (34.4%) VUS could be assigned additional pathogenic points, and 41 (5.8%) could be upgraded to likely pathogenic or pathogenic (LP/P), affecting 83 patients (Figure [H]). Of the 193 missense VUS reported in 1,496 individuals with APC data, 89 (46.1%) VUS could be assigned additional pathogenic points, and 26 (13.5%) could be upgraded to LP/P, affecting 66 patients (Figure [I]).

The addition of functional data will lower the threshold for future reclassification of a VUS, a trend recently described by Kobayashi et al. (2024)^5^. Of the 201 VUS in 550 patients that remained VUS from MAVE data and of the 63 VUS in 267 patients that remained VUS from the APC data, 121 (60.2%) and 16 (25.4%) VUS had no clinical evidence applied, respectively. This suggests that increased collaboration between clinicians and laboratories to share clinical information would facilitate further resolution of these VUS.

This study presents the impact of two distinct, but complementary functional assays, on real-world variant classification. The difference in VUS reclassification impact of 5.8% and 13.5% for MAVE and APC, respectively, is due to MAVE being an untargeted, genotype-first screening approach, while the APC data was acquired retrospectively and so has an ascertainment bias.^2^ The absolute number of reclassifications due to MAVE, however,is higher due to the increased number of variants evaluated. The NPV for both assays fell below established thresholds for awarding benign evidence. In part, this results from examining specific mechanisms of pathogenicity and not others. Additionally, the known variants used in model evaluation are heavily imbalanced towards pathogenic variants, which negatively impacts the NPV of the models. We hypothesize that as more missense benign variants are clinically classified, the NPV will improve, and reach the threshold to apply benign points to classification. Future efforts to combine the data from both assays into a single model may also improve overall performance, evidence weighting, and achieve a greater number of terminal classifications.

Our findings highlight the need for greater communication among clinicians and laboratories to share data to reclassify variants. We propose that well-calibrated functional data be used to help guide clinical laboratory VUS resolution. In the Tier 1 monogenic condition evaluated here, *KCNH2*-related LQTS, applying such functional data can immediately lend certainty to 5.8-13.5% of *KCNH2* missense variants currently classified as VUS.

## Disclosure

SRP, YT, FF, MV, SRP, and JR are current or former employees of Labcorp (formerly Invitae Corporation).

## Data Availability

All data produced in the present study are available upon reasonable request to the authors

